# NVX-CoV2373 Vaccine Efficacy Against Hospitalization: A *post hoc* analysis of the PREVENT-19 phase 3, randomized, placebo-controlled trial

**DOI:** 10.1101/2023.03.17.23287306

**Authors:** Anthony M. Marchese, Xiang Zhou, John Kinol, Eddie Underwood, Wayne Woo, Alice McGarry, Hadi Beyhaghi, Germán Áñez, Seth Toback, Lisa M. Dunkle

## Abstract

PREVENT-19, the pivotal phase 3 trial of the Novavax adjuvanted, recombinant spike protein COVID-19 vaccine (NVX-CoV2373) demonstrated that the vaccine was safe and efficacious (vaccine efficacy, VE= 90%) for the prevention of symptomatic COVID-19. In the trial, participants were randomly assigned in a 2:1 ratio to receive 2 doses of NVX-CoV2373 or placebo 21 days apart. Throughout the study, SARS-CoV-2 circulating variant was predominantly alpha, but other variants circulated (i.e., beta, gamma, epsilon, and iota). VE among the per-protocol efficacy analysis population was calculated according to pre-specified disease severity (mild, moderate, or severe) criteria, but the impact on the risk of COVID-19– associated hospitalization was not specifically investigated. During the placebo-controlled portion of the trial (January 25, 2021, to April 30, 2021), 4 hospitalizations occurred among the 77 events analyzed for the primary endpoint using the per-protocol population, 0 among vaccine recipients and 4 among placebo recipients, yielding a VE against hospitalization of 100% (95% CI: 28.8, 100). Among an expanded efficacy population, which included COVID-19–associated hospitalizations without a requirement for diagnostic polymerase chain reaction testing performed at the study central laboratory, 12 total hospitalizations were identified, 0 among vaccine recipients and 12 among placebo recipients, yielding a *post hoc* VE against hospitalization of 100% (95% CI: 83.1, 100). These additional data from the PREVENT-19 trial provide relevant public health information concerning the attributes of NVX-CoV2373.

## 1. Introduction

Since early 2020, COVID-19–associated hospitalization has placed a significant burden on the US healthcare system, requiring considerable resources, and resulting in overcrowding and other strains on hospital staff [1]. Further, it has been linked to health risks such as hospital-acquired infections [2], and substantial financial costs for individuals, families, and healthcare systems [1, 3]. The risk of hospitalization with severe acute respiratory syndrome coronavirus 2 (SARS-CoV-2) infection is understood to be heavily impacted by certain health conditions, older age, obesity, race/ethnicity, history of smoking, and occupation [4, 5], as well as social and cultural influences such as social distancing, living conditions, travel, and public mobility [6]. For COVID-19 vaccines, which have been deployed with remarkable success, the ability to prevent hospitalization is of critical importance. Despite this, vaccine efficacy estimates against hospitalization were absent from the publications associated with the pivotal trials of most widely utilized COVID-19 vaccines [7, 8].

The US-based phase 3 COVID-19 vaccine clinical trials were primarily conducted during the early pandemic stages of 2020, prior to widespread prevalence of variant strains. These trials generally reported high vaccine efficacy against symptomatic and severe disease [7, 8]. In the phase 3 trial of BNT162b2 (Pfizer/BioNTech, July 2020-November 2020), vaccine efficacy against severe COVID-19 occurrence after dose 1 was reported as 88.9% (95% CI: 20.1, 99.7), and as 75% (95% CI: -152.6, 99.5) ≥7 days after dose 2 [8]. The COVE study, a phase 3 trial of mRNA-1273 (Moderna, July 2020-October 2020), evaluated the prevention of severe COVID-19 ≥14 days after dose 2 and found a vaccine efficacy of 100% (95% CI: could not be estimated, 1.0) in the adjudicated per-protocol group, and 95.1% (95% CI: 91.1, 97.3) in a secondary case definition group [7]. Neither of these pivotal trials reported efficacy against hospitalization in the associated publications [9]. Due to widespread uptake of both mRNA vaccines, recent real-world evidence data have provided new insight into their effectiveness against hospitalization. Notably, BNT162b2 effectiveness against hospitalization from December 20, 2020, to February 1, 2021, was reported to be 87% (95%CI: 55, 100) ≥7 days after the dose 2 [10]. Other studies investigating mRNA COVID-19 vaccines effectiveness against hospitalization have found a range of estimates and are complicated by highly variable vaccination history, infection timelines, and incidental COVID-19 hospitalizations [11]. Though reductions in vaccine effectiveness against new SARS-CoV-2 variant strains (currently various omicron subvariants) have been observed, the real-world benefit of COVID-19 vaccination to reduce hospitalization rates has well been demonstrated [11-18].

The PREVENT-19 trial was the pivotal phase 3 trial of the Matrix-M–adjuvanted, recombinant spike protein COVID-19 vaccine (NVX-CoV2373), which demonstrated an efficacy of 90% for the prevention of mild, moderate, or severe COVID-19 due to circulating SARS-CoV-2 strains at the time of the trial. Although the vaccine efficacy against moderate-to-severe disease assessed by pre-defined criteria was 100%, (95% CI: 87.0, 100) the efficacy in prevention of COVID-19–associated hospitalization was not evaluated [19]. Due to the relatively short time span since the authorization of NVX-CoV2373, real-world effectiveness data that may inform its impact on hospitalization are not yet available.

This *post hoc* analysis aimed to identify all hospitalizations among the PREVENT-19 efficacy analysis population and to include additional COVID-19–associated hospitalizations that had been excluded from the per-protocol population due to absence of infection confirmation by polymerase chain reaction (PCR) testing at the central study laboratory. Further, we sought to detail clinical narratives of all hospitalized patients to better understand the underlying risk factors for COVID-19– associated hospitalization in trial participants.

## 2. Methods

### 2.1 Trial design

The methodology, protocol, demographic information, and clinical characteristics for this trial have been previously published (Dunkle et al. *NEJM* 2022; clinicaltrials.gov NCT04611802) [19]. Briefly, we assessed the safety and efficacy of two doses of NVX-CoV2373 (5 μg of SARS-CoV-2 recombinant spike protein with 50 μg of Matrix-M™ adjuvant, from Novavax, Inc. [Gaithersburg, MD]) or placebo, administered intramuscularly (deltoid muscle, using a 1” 25-gauge needle) 21 days apart (vaccine lots Par 28003 and Par 28004). This phase 3 randomized, observer-blinded, placebo-controlled trial was conducted at sites across the United States and Mexico. Participants were randomly assigned in a 2:1 ratio via block randomization to receive two doses of NVX-CoV2373 or placebo (normal saline) using a centralized Interactive Response Technology system. As part of the trial, a blinded cross-over was introduced, which ended the placebo-controlled period of the study. In the pre-crossover, placebo-controlled efficacy period endpoints accrued from January 25, 2021 to April 30, 2021. During this period 10 moderate and 4 severe COVID-19 cases were identified, all of which were among placebo recipients yielding vaccine efficacy against moderate-to-severe disease of 100% (95% CI: 87.0, 100). Disease severity was defined according to FDA-specified criteria in Dunkle et al. *NEJM* 2022. Briefly, moderate COVID-19 was defined by fever, evidence of lower respiratory tract infection (i.e., shortness of breath or abnormal chest X-ray), and/or increased respiratory rate. Severe COVID-19 case definitions included markedly abnormal vital signs, and/or requirement for high–flow oxygen therapy or mechanical ventilation. Severe case criteria also included organ system dysfunction or failure, thrombotic events, admission to the ICU, or death. Any potentially severe per-protocol endpoint cases were reviewed by an external, independent Endpoint Review Committee (iERC) to confirm principal investigator assigned severity. Neither set of severity criteria specified the need for hospitalization, and thus the specific impact of NVX-CoV2373 on hospitalizations was not reported in prior publications.

In PREVENT-19, the pre-specified analysis of efficacy within the per-protocol population required endpoint events to have a PCR positive SARS-CoV-2 infection confirmation from a nasal swab tested at the study central laboratory (University of Washington, Seattle, Washington). This protocol requirement led to the exclusion of a number of hospitalizations for COVID-19 in the primary publication [19]. To include these hospitalizations in the current analysis, vaccine efficacy was estimated with and without this requirement. All participants who received two doses of NVX-CoV2373 or placebo and were hospitalized with COVID-19 confirmed by any diagnostic assay test were included in the population hereafter referred to as the “expanded efficacy population”. Cases uniquely identified from this expanded efficacy population (those that were not part of the per-protocol efficacy analysis population) were not reviewed by the external iERC. Clinical narratives of the identified participants were obtained from the clinical study report, reviewed, and summarized.

### 2.2 Statistical analysis

To ensure datasets accurately mirrored those used in PREVENT-19 following processing and interpretation, the per-protocol efficacy analysis from the trial was confirmed **(Figure 1, Supplemental 1.1**–**1.3)**. Subsequently, two analyses with different data inclusion criteria were performed. For both, all participants in the per-protocol population (n = 25,482: NVX-CoV2373, n = 17,312; placebo, n = 8,140) within the pre-crossover date range (01/25/21 to 04/30/21) and with hospitalization events ≥7 days after dose 2 were included. In the per-protocol efficacy analysis population, participants were screened for PCR positive SARS-CoV-2 infection confirmed by the central laboratory. Among these participants all hospitalizations were identified **(Figure 1, Supplemental 1.4)**. In the expanded efficacy population analysis, the per-protocol population (n = 25,482) was used to identify participants who were hospitalized with diagnosed COVID-19 by any PCR test, rapid antigen test, or confirmed by an unspecified diagnostic assay test, irrespective of testing at the central laboratory **(Figure 1, Supplemental 1.5)**. Vaccine efficacy was defined as one minus a measure of relative risk. For both the per-protocol efficacy and expanded efficacy populations the VE and 95% confidence intervals were calculated using an exact conditional binomial method using the Clopper-Pearson method **(Supplemental Figure 1)** [20]. This method was chosen as both populations had zero events in the vaccine arm. The method involves conditioning on the total number of events across both treatment groups, where the number of events in the active group is generated from a single binomial distribution. The point estimate and corresponding confidence intervals, generated from this single binomial distribution, were then transformed back to relative risks. Data processing and interpretation were carried out using Python (Fredericksburg, VA, version 3.9.12), Pandas (Fredericksburg, VA, version 1.4.2), and Jupyter Notebook (version 6.4.8), and were recorded and described in detail. Key term abbreviations were standardized and used throughout the analysis **(Supplemental Table 1)**.

**Figure 1.**
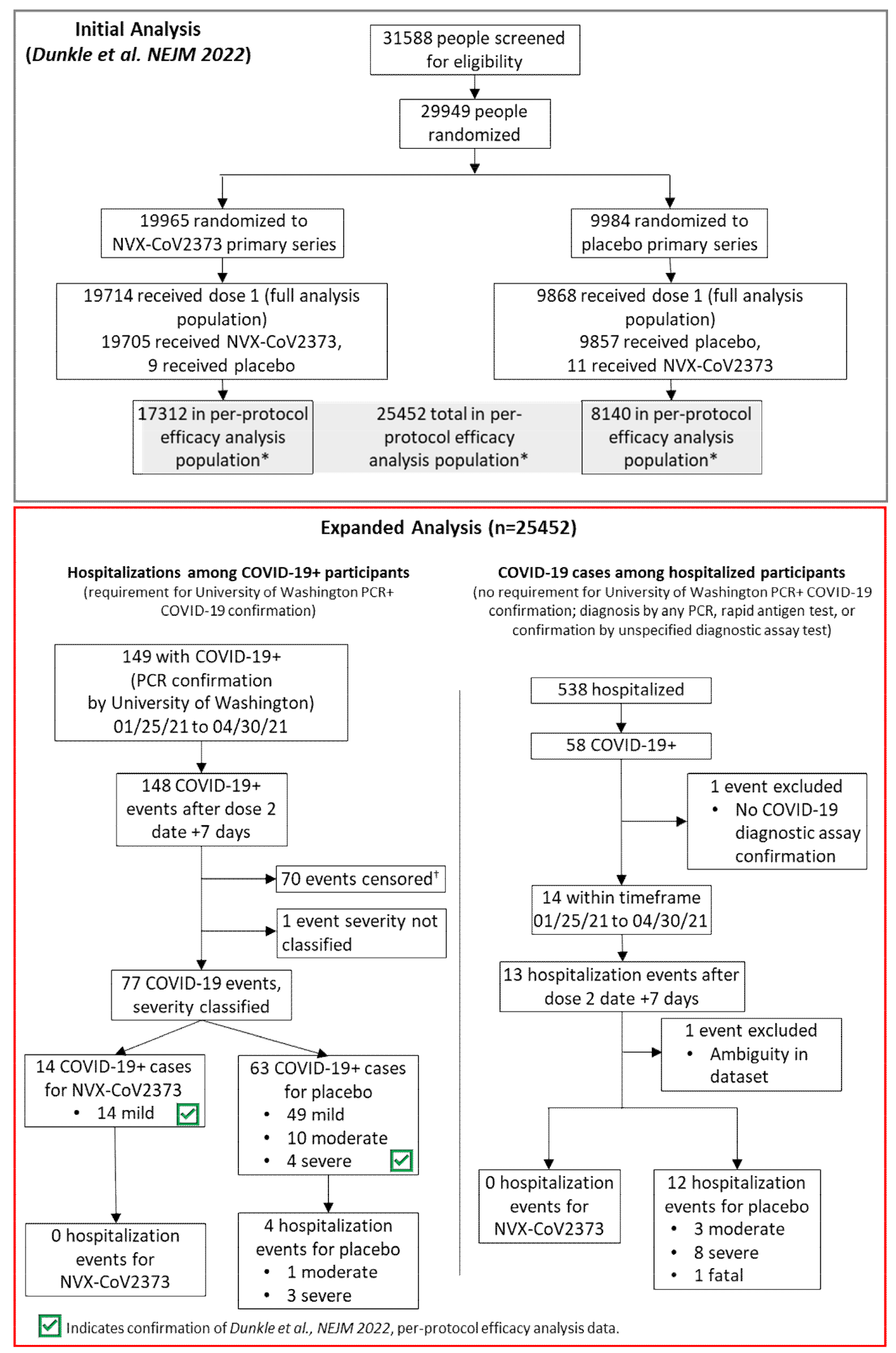
CONSORT Diagram for Original Trial and Expanded Analysis. CONSORT diagram for original trial participants and event identification in the expanded analysis. The 4 identified hospitalized participants are among the 77 events analyzed for the PREVENT-19 per-protocol primary efficacy analysis endpoint. COVID-19+ indicated diagnosis of COVID-19. *Participants received both doses as assigned, were seronegative for anti–severe acute respiratory syndrome coronavirus 2 (SARS-CoV-2) nucleocapsid protein and had a SARS-CoV-2 RNA reverse-transcriptase–polymerase-chain-reaction (RT-PCR)–negative nasal swab at baseline and did not have a censoring event at any time before 7 days after the second injection. †Censoring events for exclusion of participants from the per-protocol analysis population as described by Dunkle et al., NEJM 2022 (Supplemental S1.3).

## 3. Results

Of the 25,482 participants included in the per-protocol efficacy analysis population (NVX-CoV2373, n = 17,312; placebo, n = 8,140) 4 hospitalizations were identified where there was a PCR positive result from the central laboratory, 0 among vaccine recipients and 4 among placebo recipients **(Figure 1)**, yielding vaccine efficacy against hospitalization of 100% (95% CI: 28.8, 100) **(Figure 2)**. The 4 hospitalizations included 3 severe and 1 moderate case of COVID-19, as assessed by the iERC **(Figure 1)**.

**Figure 2.**
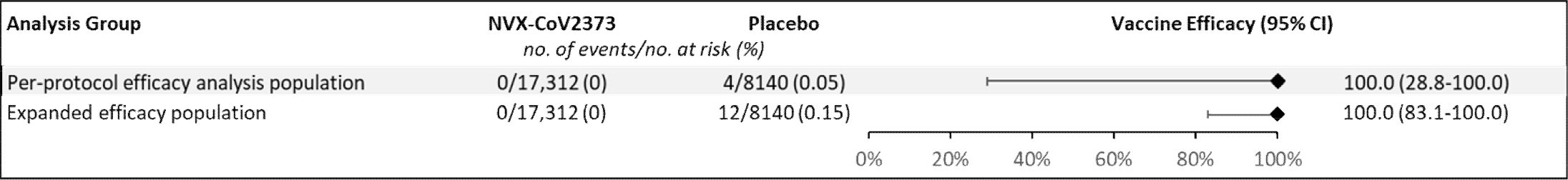
Vaccine Efficacy of NVX-CoV2373 Against COVID-19-Associated Hospitalization. Vaccine efficacy of NVX-CoV2373 against hospitalization in the per-protocol efficacy analysis and expanded efficacy populations.

With the removal of the central laboratory testing requirements pre-specified in the per-protocol efficacy analysis, 12 total (8 additional) COVID-19–associated hospitalizations were identified among the expanded efficacy population, 0 among vaccine recipients and 12 among placebo recipients **(Figure 1)**, yielding vaccine efficacy against hospitalization of 100% (95% CI: 83.1, 100) **(Figure 2)**. The 12 hospitalizations included 8 severe and 3 moderate cases of COVID-19 **(Figure 1)**.

Among the 12 identified placebo-recipients with COVID-19–associated hospitalization, 11 resolved (3 moderate cases and 8 severe cases), and 1 resulted in death. All participants were from the United States and were predominantly White or Hispanic/Latino. Participant ages ranged from 48–74 years old, with a mean (standard deviation) of 58.7 (8.2) years old **(Table 1)**. All participants had at least one pre-existing medical condition such as hypertension, hyperlipidemia, obesity, chronic obstructive pulmonary disease, gastroesophageal reflux disease and diabetes, and many were taking concurrent medications. Most participants (11, 92%) had a medical history of obesity or reported a BMI >30 kg/m^2^. Of the 9 reported BMIs, the mean (standard deviation) was 37.5 (7.6) kg/m^2^ with 6 (66%) BMIs >35 kg/m^2^ (extremely obese) **(Table 1)**. Only one participant had a BMI that did not indicate obesity, BMI=22.3 kg/m^2^. While hospitalized, most of the participants were treated with various medications, including antivirals, antibiotics, analgesics, antipyretics, and glucocorticoids, primarily dexamethasone (10, 83%), remdesivir (10, 83%), and/or enoxaparin (6, 50%) **(Table 1)**. Two of the hospitalized participants experienced sequelae suggestive of long COVID – brain fog, fatigue, shortness of breath, and loss of taste and smell – after being discharged from the hospital, and several required home oxygen therapy or cardio-pulmonary rehabilitation to regain lung function. A fatal COVID-19 case was described as severe COVID-19 pneumonia in a participant within the 55–64-year-old age range. The participant experienced shortness of breath, cough, loss of taste, chills, and fatigue for 4 days prior to being hospitalized. Despite aggressive therapeutic intervention, including bilevel positive airway pressure, broad spectrum antibiotics, albuterol, glucocorticoids, remdesivir, immune supplements, and convalescent plasma, the participant’s respiratory status continued to decline and resulted in cardiopulmonary arrest, leading to death. The participant had pre-existing medical conditions, including obesity and primary hypertension.

**Table 1.**
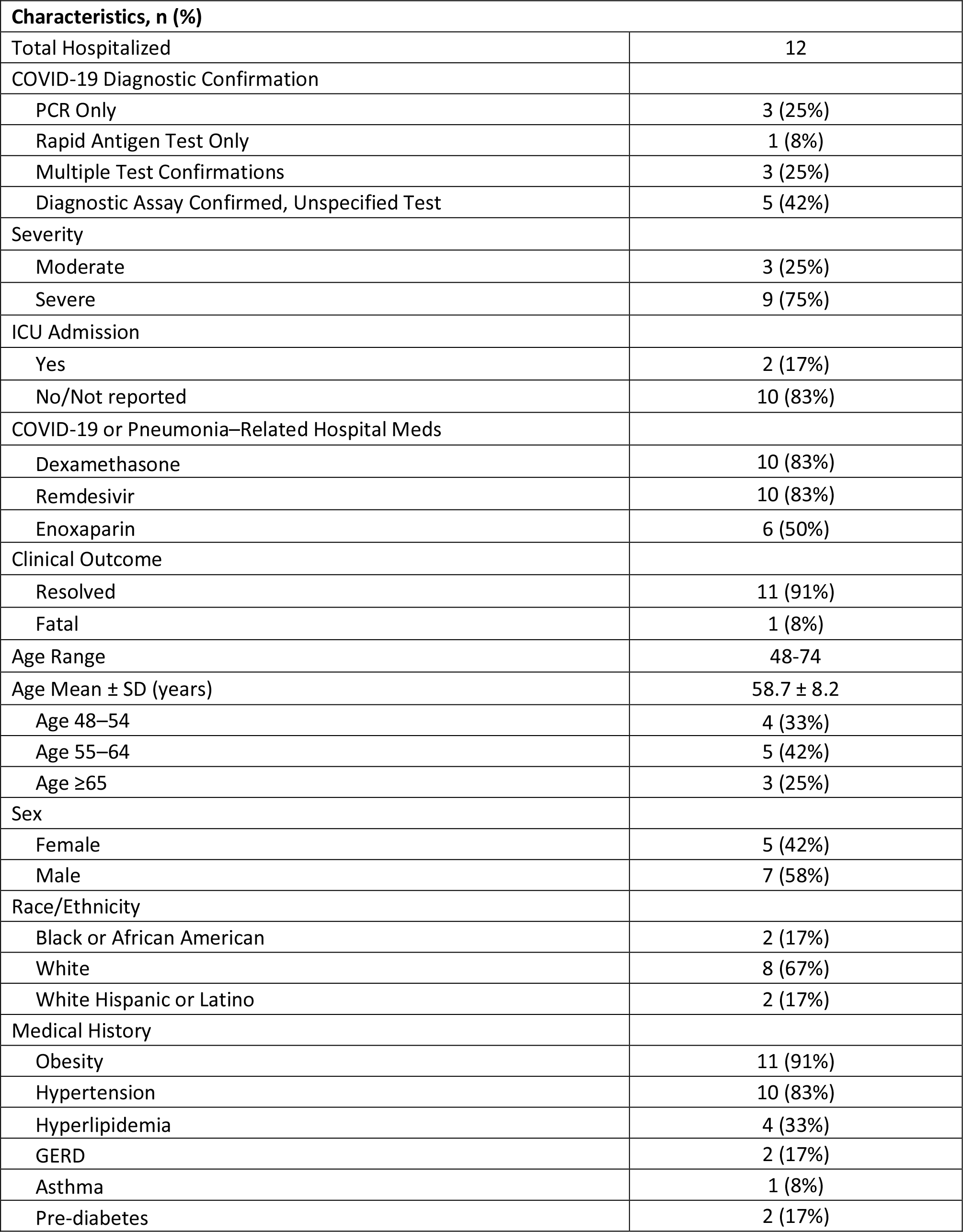
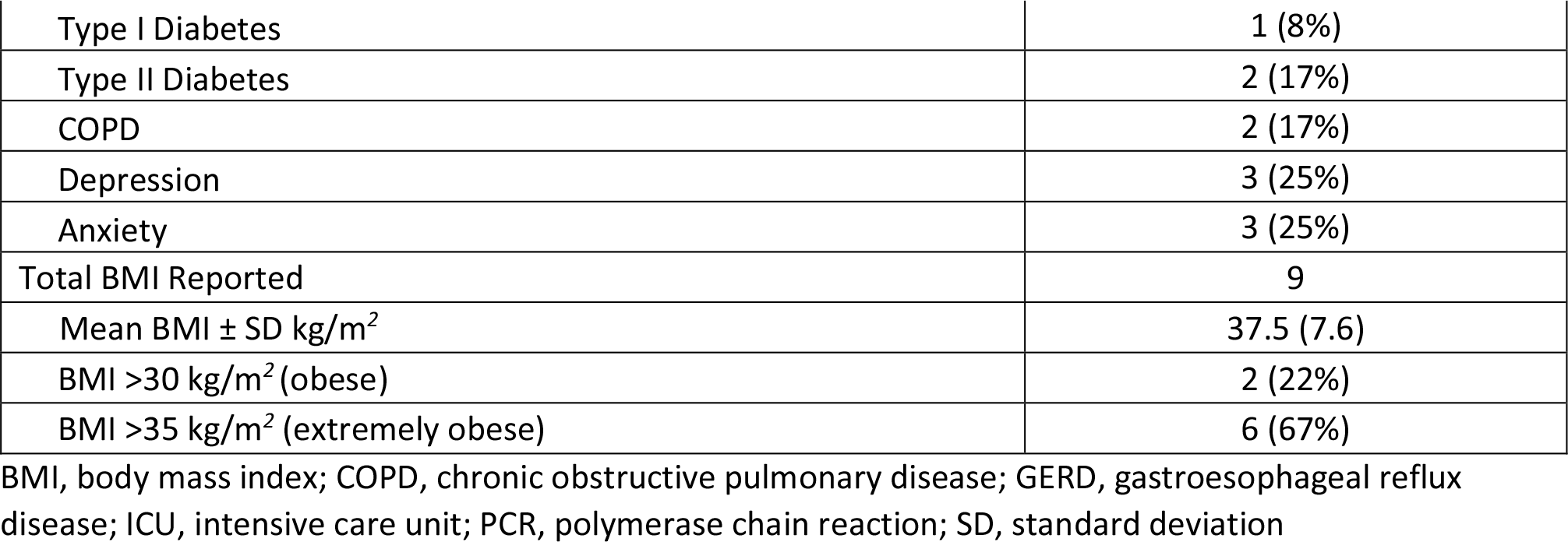
COVID-19–Associated Hospitalized Participants Among Expanded Efficacy Analysis Population

| **Characteristics, n (%)** |  |
| --- | --- |
| Total Hospitalized | 12 |
| COVID-19 Diagnostic Confirmation |  |
| PCR Only | 3 (25%) |
| Rapid Antigen Test Only | 1 (8%) |
| Multiple Test Confirmations | 3 (25%) |
| Diagnostic Assay Confirmed, Unspecified Test | 5 (42%) |
| Severity |  |
| Moderate | 3 (25%) |
| Severe | 9 (75%) |
| ICU Admission |  |
| Yes | 2 (17%) |
| No/Not reported | 10 (83%) |
| COVID-19 or Pneumonia–Related Hospital Meds |  |
| Dexamethasone | 10 (83%) |
| Remdesivir | 10 (83%) |
| Enoxaparin | 6 (50%) |
| Clinical Outcome |  |
| Resolved | 11 (91%) |
| Fatal | 1 (8%) |
| Age Range | 48-74 |
| Age Mean ± SD (years) | 58.7 ± 8.2 |
| Age 48–54 | 4 (33%) |
| Age 55–64 | 5 (42%) |
| Age ≥65 | 3 (25%) |
| Sex |  |
| Female | 5 (42%) |
| Male | 7 (58%) |
| Race/Ethnicity |  |
| Black or African American | 2 (17%) |
| White | 8 (67%) |
| White Hispanic or Latino | 2 (17%) |
| Medical History |  |
| Obesity | 11 (91%) |
| Hypertension | 10 (83%) |
| Hyperlipidemia | 4 (33%) |
| GERD | 2 (17%) |
| Asthma | 1 (8%) |
| Pre-diabetes | 2 (17%) |
| Type I Diabetes | 1 (8%) |
| Type II Diabetes | 2 (17%) |
| COPD | 2 (17%) |
| Depression | 3 (25%) |
| Anxiety | 3 (25%) |
| Total BMI Reported | 9 |
| Mean BMI ± SD kg/m<sup>2</sup> | 37.5 (7.6) |
| BMI >30 kg/m<sup>2</sup> (obese) | 2 (22%) |
| BMI >35 kg/m<sup>2</sup> (extremely obese) | 6 (67%) |

BMI, body mass index; COPD, chronic obstructive pulmonary disease; GERD, gastroesophageal reflux disease; ICU, intensive care unit; PCR, polymerase chain reaction; SD, standard deviation

One participant was excluded from the expanded efficacy population analysis due to the lack of any COVID-19 confirmation by diagnostic assay (PCR or rapid-antigen) **(Figure 1)**. Briefly, a female participant in the 48–54-year age range presented to the hospital for a pre-planned revision of her gastric sleeve and repair of a hiatal hernia with a gastroplasty approximately 25 days after receiving the second vaccination dose. During the hospitalization, she experienced sudden onset of shortness of breath which was characterized as viral pneumonia. A computed tomography scan of her chest demonstrated fibrosis due to possible pneumonitis. The participant was diagnosed with COVID-19 pneumonia based on clinical suspicion without a confirmatory COVID-19 test, and despite this, underwent gastric bypass surgery later that day. Post-operative respiratory treatments included ipratropium bromide/albuterol which were administered for 4 days. The respiratory event was considered resolved and the participant was discharged from the hospital with no additional treatment for COVID-19 pneumonia. The principal investigator (PI) assessed the event of pneumonia as severe in intensity and not related to the study vaccine. The PI did not provide an alternative etiology for the event of pneumonia. This case was not reviewed by the iERC as it was not part of the per-protocol efficacy analysis population due to a lack of PCR performed at the central study laboratory. It is important to note that the participant had a significant medical history for hypertension and her calculated BMI was 34.1 kg/m^2^.

## 4. Discussion

The pivotal clinical trials of the Pfizer/BioNTech, Moderna, and Novavax COVID-19 vaccines did not report vaccine efficacy against hospitalization, but rather focused on the evaluation of efficacy against symptomatic mild, moderate, or severe disease [7, 8, 19]. Compared with efficacy against severe disease, vaccine efficacy estimates against hospitalization can provide a clearer understanding of the vaccine’s impact on healthcare systems, thus more accurately informing public health planning and resource allocation [21]. The review of clinical narratives from vaccinated and placebo recipients captured during clinical trials can provide insightful details and a more comprehensive understanding of the impact of a vaccine on disease outcomes.

The Novavax vaccine arm showed no instances of hospitalization among participants in both the per-protocol efficacy analysis and expanded efficacy analysis populations. As a result, the vaccine demonstrated a 100% efficacy rate against hospitalization in both groups. The demographic information of nearly all identified hospitalized patients agreed with known risk factors of COVID-19 hospitalization, particularly obesity and presence of other underlying comorbid conditions including hypertension, hyperlipidemia, diabetes, chronic obstructive pulmonary disease, and psychiatric disorders [22, 23]. Compared to the per-protocol efficacy analysis population of PREVENT-19, the percentage of total participants with any coexisting conditions (47.3%) or with a medical history of obesity (37.2%) was higher among hospitalized participants from the expanded efficacy analysis population (100%, and 91%, respectively), though the latter only represents a small fraction of the former’s sample size [19]. Of hospitalized patients in the expanded efficacy population, 3 of 12 (25%) suffered from depression (compared to a US background rate of 9.5%) [24], underscoring the potential role of mental health in utilization of healthcare resources. Most hospitalized participants received COVID-19 pneumonia– related hospital medications (dexamethasone, 83%; remdesivir, 83%; and/or enoxaparin 50%). While these treatments offer great benefit to hospitalized patients, they are not without their own risks, further highlighting the importance of disease and hospitalization prevention [25-28]. Notably, one vaccinated participant presented to the hospital for an unrelated indication, though it is evident that COVID-19 infection did not lead to hospitalization due to admission for pre-planned surgery. It is plausible that this was a case of incidental COVID-19 during unrelated hospitalization, as recent research by a US-based consortium shows that incidental hospitalization, where patients are admitted for unrelated reasons and later diagnosed with COVID-19, occurs in 26% of cases [29]. In the absence of any diagnostic test to confirm a diagnosis of COVID-19, this participant was excluded from the expanded efficacy analysis population.

There are several limitations to this *post hoc* analysis. Precision of vaccine efficacy estimates from the clinical trial data is affected by the low number of events, which despite the study having approximately 30,000 participants, only included 4 COVID-19-associated hospitalizations in the per-protocol efficacy population and 12 in the expanded efficacy population. At the time of the study, pandemic era restrictions, which prevented many social events and interactions, successfully reduced infections and hospitalization rates. The trial enrollment criteria excluded certain key demographics of interest, such as those with certain underlying health conditions (i.e., some immunocompromised populations) and may not have fully reflected the general population. Here, we were able to access a more inclusive population by expanding our search beyond the per-protocol efficacy analysis population to include all participants in the expanded efficacy population within PREVENT-19 participants. The limited duration of the study window did not permit assessment of long-term vaccine efficacy, and while alpha and other early variants were prevalent in the US during the study, more information is needed to understand the impact of newer variants such as omicron. Though much information was attainable through clinical narrative reviews, these analyses were limited by what healthcare providers chose to include and may have omitted certain details such as complete medical histories. Interestingly, review of patient clinical narratives found that many hospitalized participants had common traits that may have contributed to their status. The future findings of currently ongoing real-world data and observational studies will help to provide a more complete picture of the vaccine’s effectiveness in preventing hospitalizations.

COVID-19 vaccines have played a crucial role in reducing hospitalizations by preventing the progression from mild to moderate and severe disease. Despite the impact of hospitalization on public health and economic outcomes, vaccine efficacy against hospitalization was not specifically reported in the pivotal trials of widely utilized COVID-19 vaccines. The results from this *post hoc* analysis of the PREVENT-19 study found that NVX-CoV2373 was effective in preventing hospitalization due to COVID-Nearly all hospitalized participants had pre-existing medical conditions such as hypertension, hyperlipidemia, diabetes, and obesity (BMI over 30 kg/m^2^). Importantly, the randomization scheme of PREVENT-19 resulted in well-balanced demographics between the vaccine and placebo treatment groups. Taken together, these results suggest that the vaccine is effective in reducing the risk of hospitalization due to COVID-19, especially in populations with pre-existing comorbid conditions and higher BMIs. Moving forward, real-world evidence studies on the effectiveness of NVX-CoV2373 in reducing hospitalization, particularly in high-risk populations, are needed to inform global public health policies and ensure the continued success of vaccination campaigns.

## Data Availability

Study information is available at https://clinicaltrials.gov/ct2/show/
NCT04611802, and requests will be considered

## Acknowledgements

Research, medical, and scientific support was provided by Sharon Glass of Novavax, Inc. Editorial support was provided by Kelly Cameron, PhD and Rebecca Harris, PhD of Ashfield MedComms (New York, USA), an Inizio company, supported by Novavax, Inc.

## Data Sharing Statement

Study information is available at https://clinicaltrials.gov/ct2/show/NCT04611802, and requests will be considered.

