## Supplement for "NVX-CoV2373 Vaccine Efficacy Against Hospitalization: A *post hoc* analysis of the PREVENT-19 phase 3, randomized, placebo-controlled trial"

### TABLES

Table S1: Key Term Abbreviations

| Term | Definition |
|---|---|
| ADAE | Adverse events dataset as a DF |
| ADSL | Subject level dataset as a DF |
| ADTTE | Time to events dataset as a DF |
| AESHOSP | Requires or prolongs hospitalization |
| AESTDTC | Start date/time of adverse event |
| APERIOD | Period in which adverse events happened |
| ASEV | Analysis severity/intensity |
| CESEV | Severity/intensity |
| CLASSEVT | Classification of event |
| DF | Pandas data frame |
| NUMCDOSE | Number of doses in the crossover period |
| NUMDOSE | Number of doses before the crossover period |
| NVXSEVT | Sponsor severity classification |
| PCR2MMS2 | Time to PCR confirmed mild/moderate/severe (days) |
| TRT01AN | Actual treatment for period 01 |

### S1.1 Data Processing

Three SAS datasets (same ones used for the analysis presented in Dunkle, et als, NEJM 2022) were used; (a) adsl.sas7bdat (subject level), (b) adtte.sas7bdat (time to events), (c) adae.sas7bdat (adverse events). Briefly, Python programming language and Pandas (a fast, powerful, flexible, and easy to use open-source data analysis and manipulation tool) was used to import the SAS datasets as Pandas data frames (Pandas function "read_sas"). Jupyter Notebook (a web-based interactive computing platform) was applied for programming in Python and processing data with Pandas. SAS datasets were imported as data frames, and viewed and manipulated as table-like identities (i.e. data columns are identified by a field name and data rows are identified with an index number). Each data column was transformed into a Python list for more detailed processing. The resulting analysis was held in new data frames and exported to Microsoft Excel spreadsheets with Pandas tools for additional analysis. In the following, ADSL, ADTTE, ADAE are referred to as data frames that have the subject level, time to events, and adverse events data, respectively.

### S1.2 Data interpretation

The data dictionary was used as a reference document to define the data fields of the data frames. The document "Analysis Data Reviewer's Guide - Study 2019nCoV-301" (CDISC artifact submitted to regulators as part of a data package) was referenced to provide additional context for the analysis datasets and terminology beyond that of the data dictionary.

**S1.3 Per-protocol efficacy confirmation analysis**

To ensure that the SAS datasets were imported correctly into the Python environment and that this analysis had a valid starting point, we first duplicated the results from the PREVENT-19 efficacy analysis (Dunkle et al., NEJM 2022). The following steps were preformed: (1) Filter ADSL with the per-protocol field “PPEFFFL” = “Y” to get a smaller DF adsl_pp, the length of which is the per-protocol population number: n = 25,452; (2) To confirm each arm also has the right number in adsl_pp, we applied the flag “TRT01AN” (Actual Treatment for Period 01 (N)) = 1 and 2 to get population numbers of 17,312 and 8,140 for the vaccine and placebo arms respectively; (3) The PCR-positive population within the desired time frame (01/25/21 to 04/30/21), the condition of field “PCRDT” (Date of first PCR positive) being in [01/25/21, 04/30/21] was applied to adsl_pp to get a filtered DF adsl_eff0 which has a length of 149; (4) To assure the PCRDT is after the second dose date (+7 days), we first added a V2D7 (second dose date +7 days) field, then filtered adsl_eff0 with condition of PCRDT > V2D7 to get a DF adsl_eff1 of length 148; (5) To get the subject ids from adsl_eff1, we selected the id’s from the field “SUBJID” of adsl_eff1 and define them as subjid_eff1; (6) To select the non-censoring population, we turned to ADTTE which has a data field named “CNSR”. Filtering by the condition of “SUBJID” in ADTTE being in subjid_eff1, we got a subset of ADTTE as DF adtte_eff0. Then another filtering step of “CNSR” = 0 in adtte_eff0 resulted in a DF adtte_eff1, which has a population of 78; (7) We applied the condition of “ASEV” (Analysis Severity/Intensity) being one of the [“Mild”, “Moderate”, “Severe”] values to adtte_eff1 to get a DF adtte_eff2 with a population of 77. The subject that was excluded from adtte_eff1 and not in adte_eff2 has the ASEV value of “Virologically Confirmed with greater than or equal to 1 COVID-19 disease symptom” and was not considered for analysis of the primary endpoint; (8) Each subject occurred multiple times in adtte_eff2. To get our final efficacy DF adtte_efff with unique id’s, we applied the condition of “PARAMCD" = "PCR2MMS2" in adtte_eff2. “PCR2MMS2” is the code for records with event indicator and follow-up time that are relevant to the primary endpoint, i.e., mild/moderate/severe PCR-confirmed COVID-19 occurring on or after 7 days post second dose. By doing this we ensure each subject was selected once; (9) In the final efficacy DF adtte_efff, we set “TRT01AN” = 1 to get vaccine arm DF adtte_efff _01, and “TRT01AN” = 2 to get placebo arm DF adtte_efff _02. The populations in adtte_efff _01, adtte_efff _02 are 14 and 63 respectively; (10) To get the severity of Covid-19 in the final efficacy population in each arm, we set “ASEV” = Mild, Moderate or Severe to get population numbers for each severity group respectively. For example, if adtte_efff _01 is filtered by “ASEV” = Mild, we get the mild cases for the vaccine arm; if adtte_efff _02 is filtered by “ASEV” = Moderate, we get the moderate cases for the placebo arm. The numbers in each arm and severity group are: vaccine arm: (Mild: 14, Moderate: 0, Severe: 0); placebo arm: (Mild: 49, Moderate: 10, Severe: 4).

#### S1.4 Hospitalization analysis among per protocol efficacy population

The ADAE was filtered with the condition that its “SUBJID” among the “SUBJID” of adtte_efff, we named this subset of ADAE as adae_efff. To identify hospitalized subjects, we applied the filter adae_efff with the condition of "AESHOSP" (flag for Requires or Prolongs Hospitalization) = “Y” to get a DF adae_eff_hosp. The number of subjects in adae_eff_hosp is 4 and all of them are in the placebo arm; this was obtained by setting condition “TRT01AN” = 2 in adae_eff_hosp and counting its size.

#### S1.5 Hospitalization analysis among expanded efficacy population

For hospitalization analysis beyond the 77 events included in the primary endpoint analysis, we worked with ADSL and ADAE dataset and focused attention on those adverse events that started in the same time period that was used in the per protocol efficacy analysis, i.e. (01/25/21 to 04/30/21). The detailed steps are: (1) We started with per protocol population from ADSL (“PPEFFFL” = “Y”) and joined it with ADAE, then filtered them with the conditions of "AESHOSP" = “Y” and "AEDECOD" (AE code) is either "COVID-19 pneumonia" or "COVID-19" to get a DF adae_covid_hosp of size 58; (2) We applied a time window filter to adae_covid_hosp with the condition of "AESTDTC" (Start Date/Time of Adverse Event) being between 01/25/21 and 04/30/21 to get a DF adae_covid_hosp1 with 15 participants in it; (3) To ensure that the adverse events happened after the dose 2 date + 7 days for the vaccine group in the period 1 (before the crossover), we added a date V2D7 (second dose date + 7 days) to adae_covid_hosp1, then filtered it with the condition of "AESTDTC" being later than V2D7. This resulted in a DF adae_covid_hosp2 which has a population of 14; (4) One subject (US081-0047) in adae_covid_hosp2 needed special attention. The subject received 2 doses of placebo in the period 1 and a dose of vaccine in the period 2 (crossover) on 04/21/2021. The adverse event happened on 04/28/2021, 1 week after the first dose of the active vaccine. The subject didn’t receive the second dose of vaccine in the period 2, and thus didn’t qualify for the vaccine group. The subject was excluded in the analysis because at the time of adverse event he belonged to neither the placebo group (got one vaccine shot), nor the vaccine group (didn’t get the second vaccine shot). The condition of exclusion is “not including those with (TRT0AN = 1, APERIOD = 2, NUMCDOSE = 1)” in adae_covid_hosp2, and this step resulted in a DF adae_covid_hospf with 13 subjects. “APERIOD” and “NUMCDOSE” are defined in Table 1; (5) Among the 13 subjects in adae_covid_hospf, there are 12 in the placebo group (condition of “TRT01AN” = 2 in adae_covid_hospf) and 1 in the vaccine group (condition of “TRT01AN” = 1 in adae_covid_hospf). The one in the vaccine group was excluded due to absence of any COVID-19 confirmation test in the hospital, the details are described in the results section of the main text.

**S1.6 Statistical Method for Efficacy End Points**

Figure 1: Python function for implementation of the Clopper-Pearson method.

```python
from scipy.stats import binomtest
def ve_ci(evt = 0, n = 4):
    result = binomtest(k=evt, n=n)
    r = result.proportion_ci(confidence_level=0.95, method='exact')
    ll = r[0]
    ul = r[1]
    ratio = 17312/8140.
    ve=100.*(1-(evt/(ratio*(n-evt))))
    ll_ve=100.*(1-(ul/(ratio*(1-ul))))
    ul_ve=100.*(1-(ll/(ratio*(1-ll))))
    return (ve, round(ll_ve,1),round(ul_ve,1))
```
